# Early-life enteric infection and enteropathy markers are associated with changes in adipokine, apolipoprotein and cytokine profiles later in childhood consistent with those of an adverse cardiometabolic disease risk profile in a Peruvian birth cohort

**DOI:** 10.1101/2021.03.09.21252947

**Authors:** Josh M. Colston, Yen Ting Chen, Patrick Hinson, Nhat-Lan H. Nguyen, Pablo Peñataro Yori, Maribel Paredes Olortegui, Dixner Rengifo Trigoso, Mery Siguas Salas, Richard L. Guerrant, Ruthly François, Margaret N. Kosek

## Abstract

**Background:** Metabolic syndrome is a cluster of risk factors for cardiovascular disease thought to afflict over a billion people worldwide and is increasingly being identified in younger age groups and socio-economically disadvantaged settings in the global south. Enteropathogen exposure and environmental enteropathy in infancy may lead to metabolic syndrome by disrupting the metabolic profile in a way that is detectable in cardiometabolic markers later in childhood.

**Methods:** 217 subjects previously enrolled in a birth cohort in Amazonian Peru were followed up annually from ages 2 to 5 years. Blood samples collected in later childhood were analyzed for a panel of 37 cardiometabolic biomarkers, including adipokines, apolipoproteins, cytokines, and other analytes. These were matched to extant early-life markers of enteropathy ascertained between birth and 2 years of age. Multivariate and multivariable regression models were fitted to test for associations adjusting for confounders.

**Results:** Fecal and urinary markers of intestinal permeability and inflammation (myeloperoxidase, lactulose and mannitol) measured from birth to 2 years of age were independently associated with later serum concentrations of soluble CD40-ligand, a proinflammatory cytokine correlated with adverse metabolic outcomes. Fecal myeloperoxidase was also strongly, directly associated with later levels of the anti-inflammatory adipocytokine omentin-1. Cumulative enteric protozoa exposure before 2 years of age showed stronger associations with later cardiometabolic markers than enteric viruses and bacteria and overall diarrheal episodes.

**Conclusion:** Early-life markers of enteric infection and enteropathy were associated with numerous changes in adipokine, apolipoprotein and cytokine profiles later in childhood consistent with those of an adverse cardiometabolic disease risk profile in this Peruvian birth cohort. Markers of intestinal permeability and inflammation measured in urine (lactulose, mannitol) and stool (myeloperoxidase, protozoal infections) during infancy, may predict disruptions to cytokine and adipocytokine production in later childhood that are precursors to metabolic syndrome in adulthood. Chronic enteric infections, such as by protozoan pathogens, may be more important drivers of these changes than symptomatic diarrhea or growth faltering.

**Funding:** Bill & Melinda Gates Foundation OPP1066146 and OPP1152146.

## 1. Introduction

Metabolic syndrome (MetS) refers to a cluster of risk factors for cardiovascular disease (CVD) including central obesity, hyperglycemia, hypertriglyceridemia, decreased high-density lipoprotein (HDL), and hypertension, which has long been recognized in adults and has more recently become the subject of research in children and adolescents [1]. The syndrome is thought to afflict over a billion people worldwide – three times the prevalence of diabetes – and is increasingly being identified in younger age groups, non-obese individuals and socio-economically disadvantaged settings in the global south, once thought to be protected from such conditions due to the absence of “western” lifestyle and dietary patterns [2–4]. MetS confers a twofold increased 10-year risk of CVD and a fivefold increased lifetime risk of developing type 2 diabetes mellitus (T2DM) [5], and there are complex, multidirectional mechanisms connecting each of these conditions [6]. While MetS and its sequalae can be managed and prevented through dietary, behavioral and lifestyle modifications, early identification and intervention is critical to avoid severe morbidities and the need for pharmaceutical or surgical intervention [7].

Evidence has increased over the past several decades in support of the “Barker hypothesis” that many chronic and noncommunicable conditions of adulthood have developmental origins – they are “programmed” by exposures encountered in childhood or even in utero [8–11] and MetS may be no exception. A hypothesis is forming that the etiology of this syndrome stretches into early infancy encompassing risk factors as diverse as diarrhea [10], growth [12], systemic inflammation [13] and genetic and epigenetic factors [2,14]. Such associations are challenging to characterize epidemiologically since they operate over the length of the life course; however, numerous insights have been gained through the follow-up of subjects at older ages who had previously been enrolled in birth cohort studies as infants [10,15]. For example, DeBoer and colleagues identified a statistically significant 34% increase in the odds of MetS in adulthood for every 10% increase in diarrhea burden experienced in the first six months of life by following up participants in a longitudinal study of growth and development in Guatemala some 30 years later [10]. Using similar study designs, undernutrition and sub-optimal growth in early childhood have also been shown to be associated with later life risk of CVD and glucose intolerance in both high- and lower-income countries [12].

Since the pathogenesis of MetS and its associated sequalae takes place over the life course as a result of cumulative exposures, there is an interest in identifying subclinical precursors that can be treated as biomarkers to classify individuals at elevated metabolic risk early in the disease process, facilitate early effective interventions and avert significant morbidity and organ damage [16]. This need is particularly salient given the competing diagnostic criteria for MetS, all of which rely on clinical features that occur late in the disease progression. Numerous analytes detectable in biological samples are receiving attention in this regard, and several have had correlations established, usually concurrently, with clinical cardiometabolic outcomes. Concentrations of apolipoproteins, the transporters of lipids in the circulation, can be measured in the blood, and abnormal values for these markers may precede dyslipidemia and other components of MetS [17]. Specifically, low concentrations of Apo-AI, a component of high-density lipoprotein cholesterol (HDL), are associated with poor glucose metabolism and increased risk of CVD, while apolipoprotein-B (Apo-B) is a marker of lower density lipoproteins that is more reliable than LDL, and increased levels are associated with cardiovascular risk [17,18]. The adipokines leptin and adiponectin and the ratio of the two are promising markers of MetS. Pro-inflammatory cytokines including interleukin-6 (IL-6) and tumor necrosis factor alpha (TNF-α) are directly associated with the risk of MetS, while anti-inflammatory cytokines - IL-10, ghrelin, and antioxidant factors (PON-1) – are inversely correlated with components of the cluster [16,19–22]. Several of these associations have been observed in school children and adolescents as well as adults [23,24] and mirror comparable associations with early childhood growth observed in a Peruvian cohort and elsewhere [25,26]. It is therefore possible to explore hypotheses about the early etiology of MetS using markers like these as surrogates, though there have been few published examples of this to date [13].

The objective of this analysis was to identify and characterize associations between markers of infectious enteric disease exposure and enteropathy in the first 24 months of life and adipokine, apolipoprotein and cytokine profiles in later childhood in a Peruvian birth cohort. The hypothesis was that enteropathogen exposure and environmental enteropathy (EE - chronic intestinal permeability and inflammation) in infancy disrupts the metabolic profile in a sustained and enduring manner that is already detectable in cardiometabolic markers later in childhood.

## 2. Materials and Methods

### 2.1. Conceptual framework

Figure 1 visualizes the established and hypothesized associations and causal pathways connecting early-life exposures with MetS in adolescence and adulthood that guided this analysis. For children raised in conditions of poor sanitation, environmental exposure to enteropathogens starts early in infancy and leads to EE by disrupting normal intestinal function, altering the ultrastructure and function of the delicate and complex villi and microvilli [27]. Cumulative damage to the gut’s surface increases its permeability to microbes and large molecules, provoking prolonged, low-level systemic inflammation (detectable in disruptions to cytokine profiles), and impairing uptake and altering the utilization of nutrients, which in turn leads to impaired growth and development [25]. Over the course of later childhood these processes increase adiposity and begin to irreversibly impair the body’s ability to metabolize glucose and lipids and signal insulin, eventually increasing the risk of the individual developing MetS as they progress through adolescence to adulthood [12,28]. In this analysis we tested the hypothesis that deviations from normal levels of biomarkers of inflammation, adiposity and dyslipidemia that presage later MetS are detectable in childhood and can be identified through their associations with early-life enteropathy markers.

**Figure 1:**
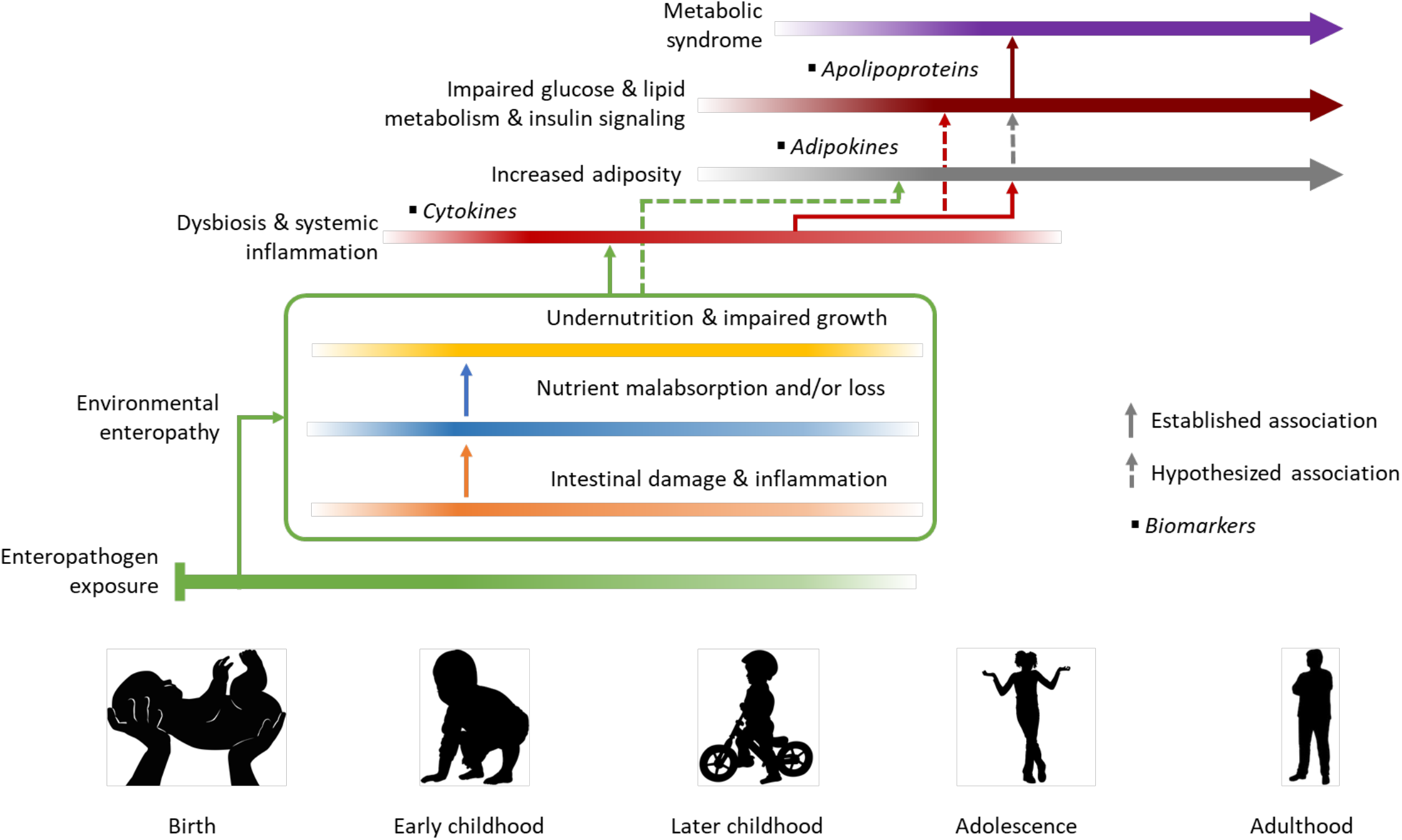
Established and hypothesized associations and causal pathways connecting early-life exposures with MetS in adolescence and adulthood and their biomarkers

### 2.2. Study population

The Etiology Risk Factors and Interactions of Enteric Infections and Malnutrition and the Consequences for Child Health and Development (MAL-ED) project was a multi-site birth cohort study that ran from November 2009 to March 2014 and was established in part to investigate the impact of EE on early childhood growth and development [29]. Subjects were recruited and enrolled (according to criteria previously described [29]) from 8 communities, each in a different Low- and Middle-Income Country and monitored continuously over their first 2 years of life through biweekly visits to generate a cumulative and continuous history of early-life disease burden. At the MAL-ED site in Peru - the village of Santa Clara de Nanay in the exurbs of Iquitos, Loreto province – contact was maintained with most of the original 303 subjects beyond the initial two-year window through weekly visits until 5 years of age. Santa Clara de Nanay is situated in a low-lying, tropical rainforest area at the confluence of several Amazon tributaries, that is characterized by a lack of infrastructure, low access to adequate water and sanitation services, and a high burden of enteropathogen infection and undernutrition [25,30]. The population of the study community are Spanish-speaking and of mixed Hispanic and Amazonian Amerindian ethnicity.

### 2.3. Outcome variables

Fasting blood samples were taken from subjects at 7 and 15 months of age and then annually from ages 2 to 5 years. The most recently contributed sample from each of 217 of the subjects was identified and analyzed using multiplex Luminex magnetic bead panels for concentrations of a panel of 37 biomarker analytes including adipokines, apolipoproteins and cytokines (Bio-Plex Pro™ RBM Metabolic and Hormone and Human Th17 Cytokine Assays; Milliplex^®^ Human Metabolic Apolipoprotein and Hormone panels). In addition, the apolipoprotein B/A-I ratio was calculated and included as a marker. For most subjects (72% of those contributing any blood samples), a sample collected at the target age of 5 years was available for inclusion, otherwise samples from the target ages of 2, 3 or 4 years (22%) or, for a minority of subjects (6%), 15 months were used.

### 2.4. Exposure variables

The following primary exposure variables were taken from extant data collected from the subjects between birth and 2 years of age as part of the MAL-ED study:

#### Biomarkers of Enteropathy

The following biomarkers were included based on their hypothesized and documented associations with EE and growth outcomes in this cohort and elsewhere [25,27,31]: seven serum biomarkers – adiponectin (ug/mL), ferritin (FRTN - ng/mL), interleukin-8 (IL-8 - pg/mL), proline, Serum Amyloid P-Component (SAP - ug/mL), serotransferrin (Transferrin - mg/dl), and tryptophan (umol/L) – measured in blood samples taken at 7, 15 and 24 months by previously described methods [25]; three fecal biomarkers—alpha-1-antitrypsin (AAT–mg/g), myeloperoxidase (MPO–ng/mL) and neopterin (NEO–nmol/L) - measured in monthly stool samples by ELISA; and the percent urinary lactulose and mannitol recovery as measured by intestinal permeability tests performed on urine samples collected at 3, 6, 9, 15 and 24 months following an oral lactulose and mannitol dosing as previously reported [32], and the lactulose:mannitol ratio was calculated and expressed as standardized as Z-scores (LMZ) relative to a low-EE reference population [33].

#### Nutritional status

Length- and weight-for-age Z scores (LAZ, WAZ) recorded at monthly anthropometric assessments were included as well as birthweight as a marker of nutritional status *in utero*.

#### Cumulative enteropathogen exposure

Stool samples collected at monthly intervals and during caregiver-reported diarrheal episodes were tested for the presence of numerous enteropathogen species using quantitative PCR, enzyme-linked immunosorbent assay (ELISA), and microscopy methods [34,35]. The pathogens with highest attributable morbidity, were summed over the two-year follow-up period for three taxa – viruses (adenovirus, astrovirus, norovirus, rotavirus, and sapovirus), bacteria (*Aeromonas* spp., *Campylobacter* spp., *Salmonella* spp., *Shigella* spp., *Plesiomonas* spp., *Vibrio* spp., and various pathotypes of *Escherichia coli*), and protozoa (*Cryptosporidium* spp., *Entamoeba histolytica*, and *Giardia* spp.). Samples from the same subject that were positive for the same pathogen were considered discrete infection episodes if separated either by an intermediate negative sample or a period of 14 days, with the exception of *Campylobacter* spp. and norovirus, for which a period of 30 days was used, and the 3 protozoa for which 3 intermediate negative samples were required (criteria previously documented by Colston and colleagues [36]). The total number of diarrhea episodes was also included.

A minimal set of covariates were used in the analysis to adjust for potential confounding including sex, household income, and age at outcome ascertainment (dichotomized at 48 months). The definitions and distributions of the exposures and covariates are presented in table 2.

#### 2.5. Statistical analysis

All biomarkers’ values were log-transformed with base 2 so that each 1 unit increase in effect size estimate was interpreted as that incurred by a doubling in biomarker concentration. Because the Luminex bead panel can only detect analyte concentrations within certain ranges of values that vary by analyte and subject age, many biomarkers had values that were above or, more commonly, below the limits of detection for the assay, and in some cases, these were a significant proportion of the overall data. Biomarkers for which more than 40% of the original values were missing were excluded from further analysis [25]. For all other biomarkers, values that were outside of the range of detection were imputed using interval regression, a method used for partially observed data with values that are missing due to being censored at known upper and lower limits, but are assumed to follow the distribution of the fully observed values [37]. For the EE biomarkers and nutritional status exposures (apart from birthweight) the within-subject average value was calculated across all available observations in the birth to 2-year age window, while for the enteropathogen infection and diarrhea episodes, the total was used to approximate cumulative exposure. These extant summary exposure measures were matched to the later outcome biomarker concentrations from the same subjects.

Correlations between the MetS serum biomarker outcomes and the early-life exposures were plotted in a correlation matrix heatmap. A multivariate regression model was then fitted to jointly model associations between the multiple early-life markers and each of the MetS biomarkers adjusting for the covariates. To assess the statistical significance of each exposure’s association with the entire set of biomarker outcomes, and vice versa, *p*-values from Wald tests of the combined significance of the relevant coefficient estimates were compared to a Bonferroni-corrected threshold of *α* = 0.05/*m* where *m* is either the number of outcomes or the number of early-life markers depending on which is being jointly tested. Finally, a subset of early-life markers was selected based on the significance and strength of their associations across multiple outcomes and included, with the same covariates, in a series of final regression models for each of the MetS biomarker outcomes. For the initial multivariate model, exposures were standardized so that their coefficients were on a similar scale and could be easily compared across multiple early-life-MetS marker pairings, whereas for the final multivariable models, they were kept on their original scale for ease of interpretation. The coefficients from the final models were visualized in a dot-and-whisker plot. Analyses were carried out using Stata 16 [38] and R 3.6.2 [39].

### 2.6. Data statement

At the time this article was submitted to the journal there were restrictions to the availability of the data used in the analysis. Readers wishing to request access to the data should contact the corresponding author

## 3. Results

Table 1 shows the definitions and categories of the variables included in the analysis. Summary statistics of the variables, including all candidate serum MetS biomarkers available for this analysis and their documented associations with individual MetS components based on a review of the published scientific literature, are presented in tables S1 and S2 in the supplementary materials. Samples from 217 subjects were analyzed for biomarkers including adipokines, apolipoproteins, cytokines, hormones, peptides, proteins, and an enzyme. Prior to applying the exclusion criteria, 37 distinct MetS biomarkers were initially considered for inclusion in further analysis. Fourteen analytes - amylin, ghrelin, glucagon-like peptide 1 (GLP-1), glucagon, interferon gamma (IFN-γ), interleukins 1β, 4, 6, 10, 17A, 22, 23 and 33, and Peptide Tyrosine Tyrosine (TYY) - were excluded from further analysis due to more than 40% of their original values being missing (mostly due to their concentrations being below the quantifiable range for the assay). The 23 analytes that met the inclusion criteria of retaining more than 60% of their original values included all of the apolipoproteins (Apo), the adipokines C-peptide, Fibroblast Growth Factors 21 and 23 (FGF21 and FGF23), Galectin-3 (Gal-3), Gastric Inhibitory Polypeptide (GIP), insulin, leptin, omentin-1, Pentraxin 3 (PTX3), Paraoxonase-1 (PON1), Pancreatic Polypeptide (PP), Soluble Leptin Receptor (sOB-R), and Visceral Adipose Tissue-Derived Serpin (Vaspin), and the cytokines Monocyte Chemoattractant Protein-1 (MCP-1), Soluble CD40-ligand (sCD40L), and Tumor Necrosis Factor Alpha (TNF-α). Of these, 5 (omentin-1, ApoA-I, PON1, sOB-R and PP) have a documented inverse association with MetS components overall, while the remaining 18 had direct associations previously reported in the literature.

**Table 1:**
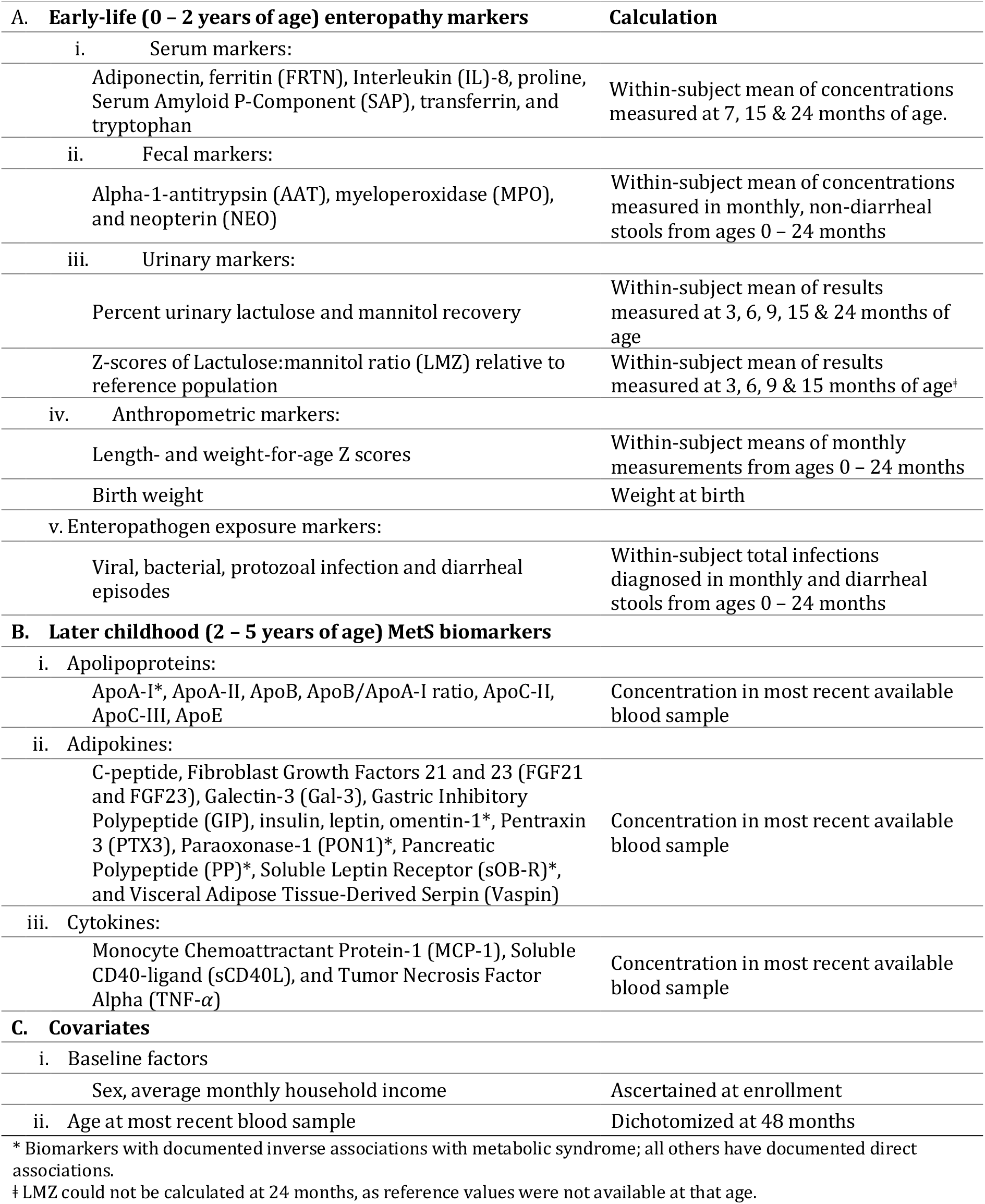
Definitions of early-life enteropathy markers, later childhood MetS markers that met the criteria for inclusion in the analysis, and covariates.

Figure 2 shows a heatmap matrix of the Spearman coefficients for the correlations between each of the included serum MetS biomarkers with each other and with the early-life enteropathy markers. As expected, correlations between markers in early-life exposures with those measured later in childhood tended to be weaker than those between the later life biomarkers and each other. Notable exposure-outcome correlations include that of fecal MPO concentration, an acute intestinal inflammation marker, directly with omentin-1 and inversely with sCD40L, as well as WAZ-score with leptin. Among the correlations between MetS markers, the apolipoproteins demonstrated strong direct correlations exclusively with each other. Other strong correlations included C-peptide directly with both GIP and insulin and the inverse correlation of omentin-1 with sCD40L.

**Figure 2:**
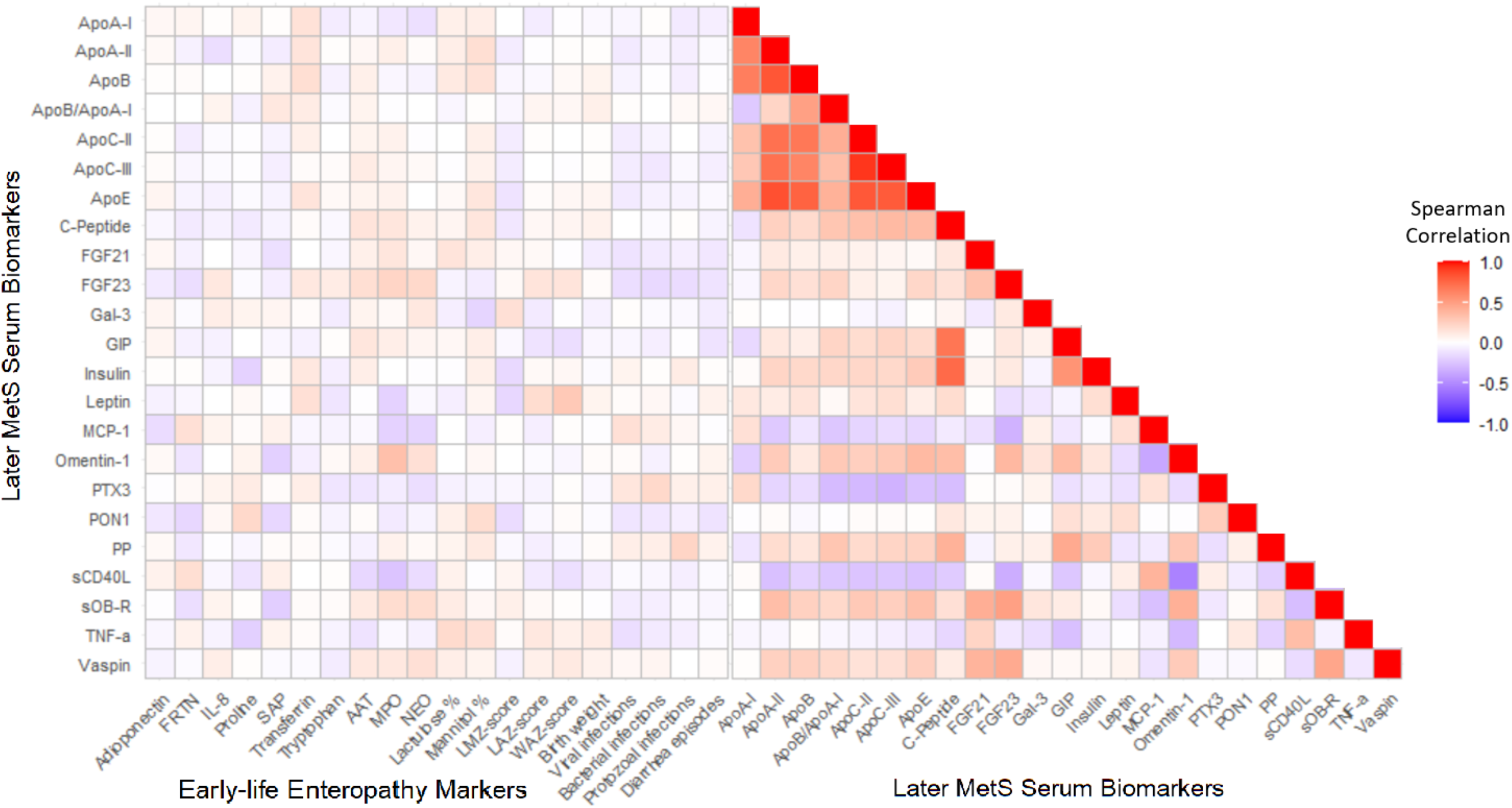
Heatmap matrix of the correlations between each of the included serum MetS biomarkers with each other and the early-life enteropathy markers

The heatmap matrix in figure 3 shows the coefficient estimates from a multivariate linear regression model of the associations between the set of standardized early-life markers and the cardiometabolic biomarker outcomes in late childhood adjusting for the covariates sex, income, and age at follow-up. The serum biomarkers are ordered along the y-axis according to the strength and direction of their documented association with MetS based on the review of the literature. Multiple early-life exposure markers showed strong or statistically significant associations with the biomarker outcomes measured in later childhood. By far the largest effect sizes observed were the direct association of urinary lactulose with sCD40L, and the corresponding inverse association between urinary mannitol with that same cytokine. For numerous other MetS biomarkers, the effects for lactulose compared to mannitol were similarly in opposing directions, while for the LMZ-score (calculated from the two urinary biomarkers, but unavailable for 24-month samples due to lack of standard reference values at that age), no strong or statistically significant effects were observed for any MetS biomarker. sCD40L was also strongly inversely associated with IL-8, proline, MPO, and protozoal infections and was statistically significant at the Bonferroni-adjusted *α* level across the entire set of exposures. Other outcomes that were statistically significant by the same criterion included TNF-*α* and omentin-1, respectively the biomarkers with the strongest direct and the strongest inverse associations with MetS documented in the literature by our assessment, as well as leptin. The most statistically significant effect of any single early-life-later childhood marker pairing was the direct association between MPO and omentin-1. While MPO also showed strong inverse associations with MCP-1 as well as sCD40L and leptin and a direct effect on FGF21, the marker did not retain statistical significance at the Bonferroni-adjusted *α* level across the entire set of MetS biomarkers. Proline, transferrin, and total protozoal infections were the only early-life markers that met this criterion of correction for multiple comparisons, the latter showing strong inverse associations with MCP-1, sCD40L, leptin and ApoA-I, and direct associations with insulin, PP and sOB-R.

**Figure 3:**
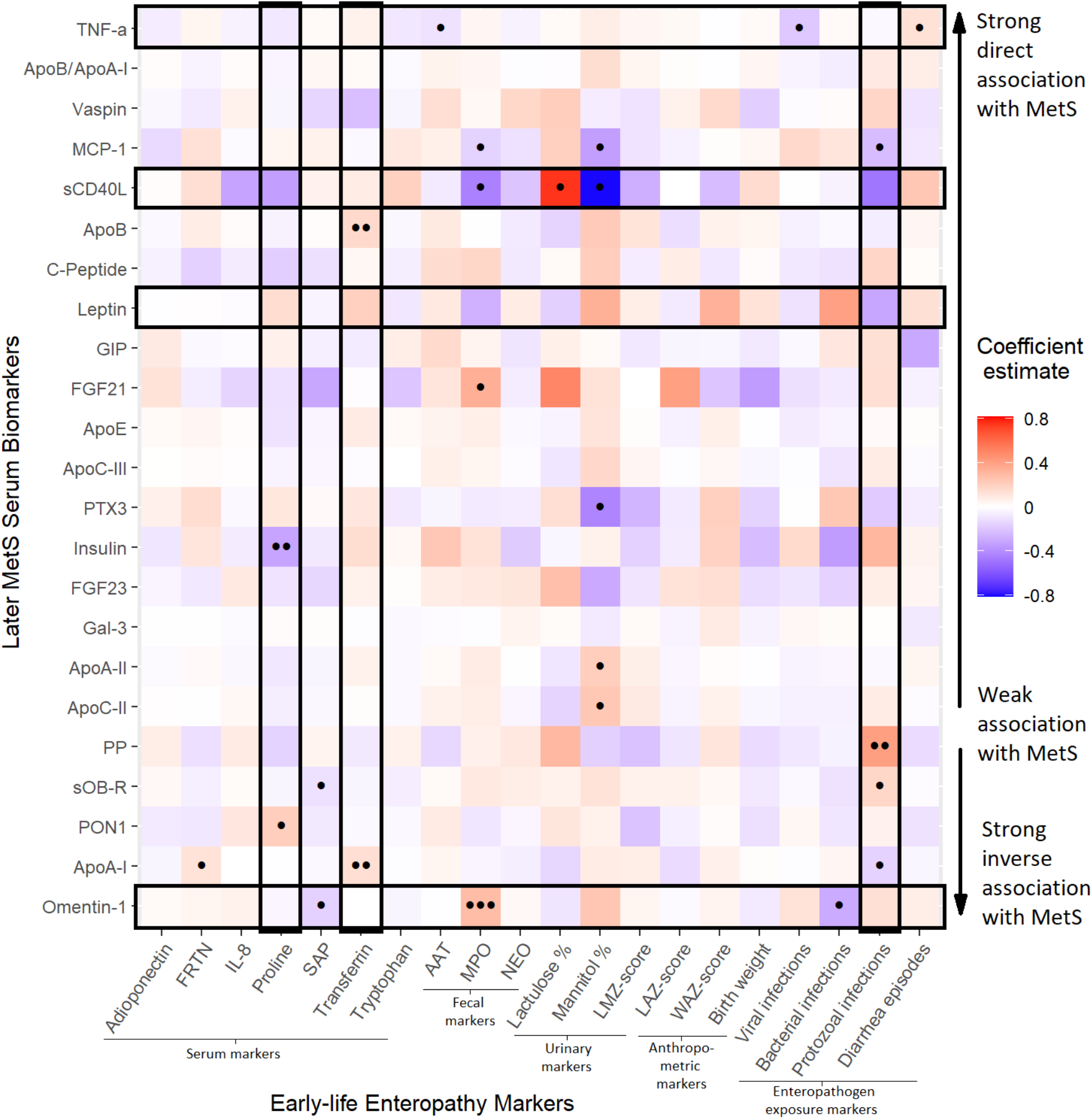
Heatmap matrix of the coefficients from a multivariate regression model of the associations of multiple, standardized early-life enteropathy markers with MetS biomarkers later in childhood – ordered by the strength and direction of their documented association with metabolic syndrome (MetS) - adjusting for sex, income, and age at follow-up. Unadjusted significance levels for individual coefficients are indicated with dots (••• *p*<0.001, •• *p*=0.001-0.01, • *p*=0.01-0.05) and combinations of outcomes and exposures that were significant at the Bonferroni-adjusted α ≤ 0.05/*m* level according to the Wald test are outlined.

The subset of six early-life markers selected for inclusion in the final regression models consisted of fecal MPO, percent urinary lactulose and mannitol recovery, serum proline and transferrin and total protozoal infections, the latter rescaled so that the coefficients represent the effects of a 10-infection increment. Figure 4 plots the coefficient estimates from the models fitted for the subset of early-life markers on each of the log_2_ MetS biomarker concentrations adjusting for the covariates. As in the multivariate model, the effects of lactulose and mannitol on sCD40L were among the largest identified (though no longer the largest overall) and almost equivalent in magnitude but in opposite directions, a doubling in lactulose recovery associated with a more than two thirds increase (0.68 [0.08, 1.28]) and the equivalent increase in mannitol with a similarly sized decrease (0.59 [-1.17, −0.02]) in sCD40L concentrations in later childhood. MPO and proline concentrations in infancy also had large inverse effects on later sCD40L levels – respectively a decrease of 1.08 (−1.70, −0.46, the largest effect size identified by the final models) and 0.75 (−1.47, −0.03) in log_2_ concentrations for each one-unit increase – as did total protozoal infections, though in the latter case the confidence intervals for the estimate narrowly spanned the null value.

**Figure 4:**
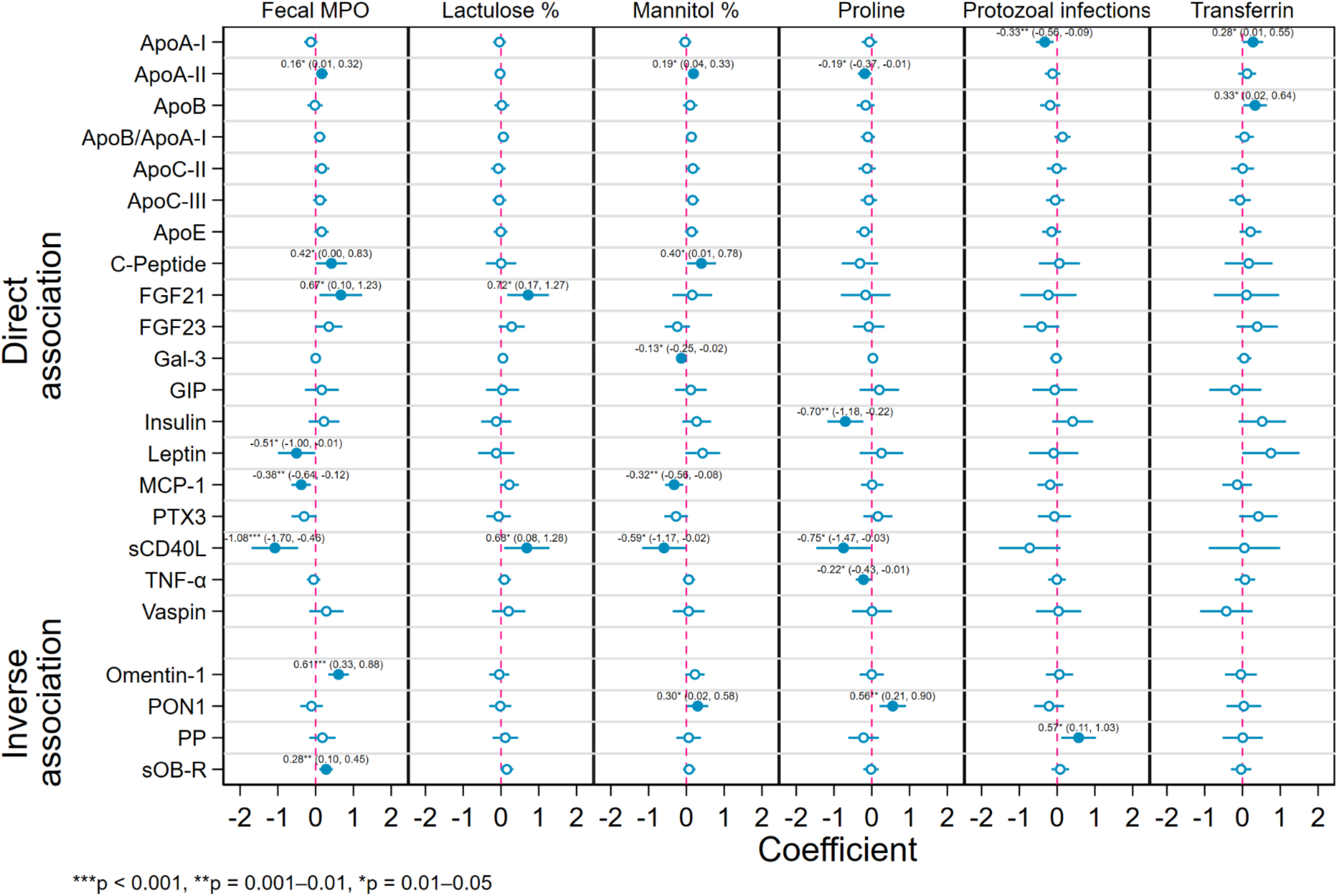
Coefficient estimates from regression models fitted for a subset of six early-life exposures – myeloperoxidase (MPO), percent urinary lactulose and mannitol recovery, serum proline and transferrin and protozoal infections (scaled so the effect is for an increase in 10 infections) - on each of the cardiometabolic biomarker outcomes adjusting for sex, income, and age with 95 % confidence intervals and significance levels. Outcomes are grouped by the documented direction of their associations with metabolic syndrome (direct or inverse).

The effect of early-life MPO on later omentin-1 levels remained the most statistically significant association identified in the final models. An increase in omentin-1, concentrations of almost two thirds (0.61 [0.33, 0.88], *p* = 0.00002) was identified for every doubling in MPO levels, though none of the other 5 enteropathy markers in the final subset showed significant effects on omentin-1. MPO also showed large direct effects on FGF21 and C-Peptide and smaller effects on sOB-R and ApoA-II as well as indirect effects on sCD40L, leptin and MCP-1. In addition to sCD40L, mannitol showed smaller-magnitude inverse associations with MCP-1, Gal-3 and PTX3 (the latter not significant at the *α* = 0.05 level), and small direct associations with ApoA-II, C-Peptide and PON1, while for lactulose the only other biomarker to show a statistically significant association apart from sCD40L was FGF21, the larger of the two effect sizes (0.72 [0.17, 1.27]). Serum proline concentration measured in early life demonstrated inverse associations with several later-life biomarkers that are directly associated with MetS, notably insulin (−0.70 [-1.18, −0.22]) and sCD40L (−0.75 [-1.47, −0.01]) – and a direct association with PON1 (0.56 [0.11, 1.03]), which is itself directly associated with MetS.

An increase in 10 total protozoal infections between ages 0-2 years was associated with a more than 50% increase in PP (0.57 [0.11, 1.03]) and a one-third decrease in ApoA-I (−0.33 [-0.59, −0.09]), as well as a notable increase in insulin concentration and decreases in sCD40L and FGF23, though the latter three with confidence intervals that narrowly included the null value. Although it was one of just two early-life markers to be statistically significant at the Bonferroni-adjusted α −level in the initial multivariate regression, transferrin only showed associations with ApoB (0.33 [0.02, 0.64]) and ApoA-I (0.28 [0.01, 0.55]) that were significant at the at the α = 0.05 level in the final models, though large direct effects on leptin, insulin and PTX3 and an indirect effect on vaspin were also observed. Similarly, though TNF-α had been significantly associated with the full early-life marker panel in the multivariate model, it only showed a small, statistically significant association with proline in the final subset (−0.22 [-0.43, −0.01]).

## 4. Discussion

There is an urgent need to clarify the mechanisms that link enteric infections and EE in infancy with MetS later in life in a way that can inform intervention early in the disease process before significant end-organ damage and morbidity occur [16]. It is plausible that the documented associations between diarrhea in early infancy, undernutrition in early childhood, and MetS later in life are partially mediated by EE. Further evidence for this hypothesis comes from the observations that MetS is highly prevalent even in low-resource settings with high enteropathogen burdens and an absence of lifestyle risk factors, and that obesity and liver steatosis are associated with concurrent intestinal permeability in adults [40,41]. Hypothesized pathways by which EE in infancy may be linked to subsequent MetS in childhood and beyond include depressed apolipoprotein production causing dyslipidemia and inflammation due to bacterial or lipopolysaccharide translocation [13]. However, the magnitude and relative clinical significance of these pathways have yet to be characterized. This study represents an initial attempt to investigate the associations between markers of early-life environmental exposures and adipokine, apolipoprotein and cytokine profiles later in childhood. It is also the first to report the distributions of this large panel of biomarkers in a cohort of young children.

According to our conceptual framework the earliest metabolic alterations may manifest as disruptions to cytokine profiles. While many cytokines that were tested for in this cohort could not be included due to having too many values outside the detectable range (e.g., IFN-γ and various interleukins), three that were included (MCP-1, sCD40L and TNF-α) demonstrated notable associations with numerous early-life enteropathy markers. In particular, concentrations of sCD40L, a proinflammatory cytokine shown to be positively correlated with concurrent insulin resistance, BMI and waist circumference [42,43], was strongly associated with lactulose and mannitol in opposite directions but not with Z-scores of the L:M ratio, the more commonly used way of expressing these markers as an indicator of intestinal absorption and permeability [44]. The L:M Z-scores did not include data from 24-months due to the lack of available standard values at that age, which may contribute to the difference in effect compared with the two component biomarkers individually. As a larger, disaccharide molecule, lactulose is absorbed in greater proportions when permeability is high due to mucosal cell damage, while the monosaccharide mannitol is instead absorbed in direct proportion to the healthy mucosal absorptive area [32,44]. Consequently, inflammatory bowel syndromes (such as Crohn’s disease) are associated with higher excretion of urinary lactulose, conditions characterized by flattening of the villi (e.g., Celiac disease) with lower mannitol [45], and both enteropathies are associated with elevated sCD40L levels [46,47]. This analysis also identified similar inverse associations between mannitol and both PTX3 and MCP-1, which, like sCD40L, are inflammatory markers themselves correlated with adverse MetS outcomes [48,49].

Potential biomarkers of increased adiposity that were available for this analysis include the pro-inflammatory adipokines leptin, vaspin, FGF21, and TNF-*α* (the latter also a cytokine), which are known to increase as a result of the interacting effect of central obesity with the other four MetS components (hyperglycemia, hypertriglyceridemia, dyslipidemia, and hypertension) [21]. The direct association between diarrhea episodes and TNF-α in the multivariate analysis may constitute further evidence that previously documented links between diarrhea in early childhood and MetS in adulthood may be mediated by increased adiposity with EE as its etiology [14]. Specifically, EE may disrupt metabolic function through pathways other than increased intestinal permeability, such as through increased adiposity as a result of elevated energy harvest caused by gut dysbiosis [28]. However, the strong effect of both urinary lactulose and fecal MPO on FGF21, an adipokine associated with increased intestinal permeability in mice and obesity in humans, suggests a complex interrelationship between these risk factors in the early pathogenesis of MetS [50,51].

As biomarkers of lipid metabolism, we hypothesize that disruptions to apolipoproteins become apparent later in the MetS pathogenesis than for cytokines and adipokines (figure 1), accompanying later stage adverse metabolic adaptations [52,53]. Nevertheless, some small-magnitude associations with early-life exposures were observed here even for apolipoproteins measured in childhood, though only for ApoA-I (directly with transferrin, inversely with protozoal infections), ApoA-II (directly with MPO and mannitol, inversely with proline), ApoB (directly with transferrin), and ApoC-II (directly with mannitol). Notably, the ApoB/ApoA-I ratio, thought to be a better predictor of insulin resistance and MetS than any single lipid fractions [53], showed no associations with early-life markers, while for its two component analytes several associations in the same direction were observed. These simultaneous positive associations with transferrin, which is directly correlated with infant growth in this cohort [25], suggest that both ApoA-I and ApoB in children from EE-endemic settings may predict nutritional status, since loss of intestinal surface area is associated with decreased levels of both apolipoproteins [13].

While these results appear consistent with the conceptual framework outlined in figure 1, others appear to be at odds with prior hypotheses and findings. Fecal MPO - a lysosomal protein found in neutrophils that is used as a marker of oxidative stress induced by inflammatory bowel disease (IBD) and EE and is predictive of subsequent growth faltering in young children [25,27,54,55] – had an inverse association with sCD40L that was larger and more statistically significant than that of mannitol, as did proline, another documented predictor of poor growth and oxidative damage [25,56]. Previous findings have shown plasma sCD40L to be positively correlated with concurrent levels of inflammatory chemokines [57]; so, it is unclear why IL-8, as well as oxidative stress markers (MPO and proline), would be negatively associated with later levels of the cytokine in this study. Furthermore, the most statistically significant association found here was the direct effect of MPO on omentin-1, an anti-inflammatory adipocytokine known for being inversely correlated with weight loss, beneficial for insulin-sensitizing and vasodilation and protective against metabolic and other disorders [58,59]. However, it is now understood that omentin-1 is also an acute-phase, anti-inflammatory reactant, and therefore may increase in the initial stages of disease and as a response to inflammation [59]. Omentin-1 also has a protective role against oxidative stress, such as is produced by MPO-induced cell injury [60], which may explain the positive association between these two markers in our analysis. Since oxidative stress is closely implicated in the pathogenesis of atherosclerosis in MetS, assessing both markers in combination may be clinically relevant and necessary for predicting adverse health outcomes. This is consistent with previous findings that intestinal T regulatory cells are activated and more abundant in children with active IBD than healthy controls [61].

Previous longitudinal studies have shown a link between poor early childhood growth and increased incidence of CVD [62] and glucose intolerance [63] later in life. The fact that significant associations were observed for biochemical enteropathy markers but not for the anthropometric indicators in this study suggests that EE may impact later metabolic outcomes via mechanisms that operate independently of nutritional status and for which growth is a marker, rather than a mediator. In contrast to previously published findings, we did not find strong associations between diarrhea episodes in early childhood and later MetS markers [10] and of the three pathogen taxa assessed, only cumulative protozoal infections showed substantial effects on the later biomarkers and mostly only in the multivariate model, such as increases in insulin, PP and sOB-R and decreases in leptin, MCP-1, sCD40L, and ApoA-I concentrations. sCD40L protects against protozoal infections through its interaction with CD40 promoting cell-mediated immunity [64], while leptin mediates resistance to *E. histolytica* [65]; so, it is possible that these infections are acting as a marker of chronic deficiency in these two cytokines that lasts through childhood. However, leptin is secreted by colonic epithelial cells during acute inflammation and both micro and macro parasitic intestinal infections can disrupt its production [66–68], so, a causative relationship is also plausible. Leptin and its binding protein sOB-R are reciprocally regulated [69], explaining these two analytes’ inverse correlation and opposing associations with protozoal infections in this cohort and with MetS in previous literature. The chemokine MCP-1 is involved in the intestinal inflammatory immune response against infection with various pathogens and in this study, we demonstrate an increase in enteric protozoal infections associated with lower serum MCP-1 for the first time in a human population (figure 3) [70]. While this apparent suppression of pro-inflammatory cytokines MCP1 and sCD40L due to protozoal infections may at first sight seem surprising, previous studies have documented diminished inflammatory immune responses following repeated intestinal parasite infections, perhaps as an immune tolerance response to prevent further tissue damage [71]. Such a downmodulation mechanism may also underlie the equally surprising inverse associations of IL-8, MPO and proline with sCD40L. The direct association between cumulative protozoal infections and PP suggests that the loss of appetite that is a common symptom of those infections may be mediated, as opposed to compensated for, by the appetite suppressing peptide [72]. The strong direct association between protozoal infections and blood insulin levels appears to have no precedent in prior literature. A possible explanation for protozoal infections showing more associations with later outcomes than viruses and bacteria may be that they are more prone to longer, chronic infections, suggesting that cumulative duration of infection may be more important than the number of discrete infection episodes.

This analysis was subject to several limitations. Not only were numerous cytokines that circulate at very low concentrations not able to be quantified in the cohort, but other biomarkers of documented relevance were also unavailable, such as the adiponectin-leptin ratio [73], since adiponectin was only measured in the early-life assessments, and leptin at outcome ascertainment. Furthermore, as with any cohort study, the possibility that those subjects retained under surveillance differed systematically from those lost to follow-up is a potential source of bias that should be considered when interpreting these findings. The principal limitation, however, is the relatively short interval that elapsed between ascertainment of the exposures and outcomes. Although MetS is increasingly recognized as manifesting in childhood, the subjects in this study may have still been too young at the time of follow-up (≤5 years) for the precursors of the conditions to have become fully apparent. This may explain why many of the effect sizes identified here were small in magnitude; however, the fact that they were apparent at all and statistically significant even in mid-childhood is itself striking. We anticipate that these divergences in metabolic profiles will continue into adolescence and beyond and be accelerated by puberty and the accompanying increases in insulin, growth hormone and IGF-I secretion, changes in body composition and slowing of metabolism [74]. This highlights the need for longer follow-up at multiple stages of the life course in this and other cohorts.

## 5. Conclusions

In conclusion, early-life enteric infection and enteropathy markers are associated with numerous changes in adipokine, apolipoprotein and cytokine profiles later in childhood consistent with those of an adverse cardiometabolic disease risk profile in this Peruvian birth cohort. In particular, markers of intestinal permeability and inflammation measured in urine (lactulose, mannitol) and stool (MPO, protozoal infections) during infancy, may predict disruptions to cytokine and adipocytokine production in later childhood that are precursors to MetS in adulthood. Chronic or recurring enteric infections, such as by protozoan pathogens, may be more important drivers of these changes than symptomatic diarrhea or growth faltering.

## Data Availability

Readers wishing to request access to the data should contact the corresponding author after publication.

## 6. Acknowledgements

We wish to thank participants, their families and the study community for their dedicated time and effort to better the understanding the transmission and more enduring impact of enteric infections is early childhood.

## 7. Author contributions

**Josh Colston:** Conceptualization, methodology, formal analysis, writing – original draft, visualization, supervision. **Yen Ting Chen:** Formal analysis, investigation, writing – original draft. **Patrick Hinson:** Formal analysis, visualization. **Nhat-Lan Nguyen:** Writing – original draft. **Pablo Yori:** Investigation, supervision. **Maribel Olortegui:** Investigation, supervision. **Dixner Trigoso:** Investigation. **Mery Salas:** Investigation. **Richard Guerrant:** Conceptualization, writing – review and editing. **Ruthly Francois:** Investigation, writing – review and editing. **Margaret Kosek:** Conceptualization methodology, writing – review and editing, supervision, funding acquisition.

## 10. Ethical standards

Ethical approval for MAL-ED was given by the Johns Hopkins Institutional Review Board as well as the Ethics Committee of Asociacion Benefica PRISMA, and the Regional Health Department of Loreto;

