## Supplementary Materials for "Early-life enteric infection and enteropathy markers are associated with changes in adipokine, apolipoprotein and cytokine profiles later in childhood consistent with those of an adverse cardiometabolic disease risk profile in a Peruvian birth cohort"

**Table S1: Summary statistics of candidate outcome biomarkers of metabolic syndrome (MetS) in the Peruvian birth cohort and their documented association with MetS components[1]**

| Analyte | Mean | Std. Dev. | Min. | Max. | Available values (%) | Associations with MetS components | Reference |
|---|---|---|---|---|---|---|---|
| **Amylin (total)** | 22.6 | 32.8 | 0.9 | 394.4 | 100 (46.1) | TG+, IR+, BMI+ | (1) |
| **Apolipoprotein A-I (ApoA-I)** | 976,625.3 | 486,208.5 | 181,996.7 | 3,423,800.0 | 217 (100.0) | HDL+, TG-, IR-, WC-, BP- | (2) |
| **Apolipoprotein A-II (ApoA-II)** | 368,648.9 | 142,495.6 | 73,415.9 | 1,268,400.0 | 217 (100.0) | WC+ | (3) |
| **Apolipoprotein B (ApoB)** | 1,514,265.8 | 734,005.1 | 236,557.0 | 4,335,500.0 | 217 (100.0) | HDL-, TG+, IR+, WC+, BP+ | (2) |
| **ApoB/ApoA-I ratio** | 0.6 | 0.6 | -1.5 | 2.0 | 217 (100.0) | HDL-, TG+, IR+, WC+, BP+ | (2) |
| **Apolipoprotein C-II (ApoC-II)** | 178,873.2 | 85,068.6 | 30,553.7 | 512,836.1 | 217 (100.0) | TG+ | (4) |
| **Apolipoprotein C-III (ApoC-III)** | 309,030.6 | 124,960.6 | 54,546.3 | 849,204.9 | 215 (99.1) | TG+, IR+, BP+, LDL+ | (5) |
| **Apolipoprotein E (ApoE)** | 76,452.0 | 36,270.9 | 15,387.6 | 221,063.7 | 217 (100.0) | TG+, IR+, BMI+ | (6) |
| **C-Peptide** | 787.6 | 627.6 | 19.9 | 2,683.4 | 210 (96.8) | HDL-, TG+, IR+, BMI+ | (7) |
| **Fibroblast growth factor 21 (FGF21)** | 324.1 | 482.4 | 1.2 | 3,008.8 | 203 (93.5) | HDL-, TG+, BMI+, WC+ | (8) |
| **Fibroblast growth factor 23 (FGF23)** | 278.3 | 423.7 | 2.0 | 3,271.2 | 216 (99.5) | HDL-, TG+, BMI+, WC+ | (9) |
| **Galectin-3 (Gal-3)** | 21,483.1 | 7,218.3 | 4,745.9 | 63,844.5 | 197 (90.8) | IR+, BMI+ | (10) |
| **Ghrelin** | 6.2 | 30.8 | 0.0 | 437.1 | 15 (6.9) | HDL+, IR-, BMI-, BP- | (11) |

[1] +, direct association; -, inverse association; TG, triglycerides; IR, insulin resistance; BMI, body mass index/obesity; WC, waist circumference/central adiposity; BP, blood pressure; HDL, high-density lipoprotein cholesterol; LDL, low-density lipoprotein cholesterol. All analyte concentrations are in units of pg/mL, except for the apolipoproteins which are in ng/mL.

**Table S1: Summary statistics of candidate outcome biomarkers of metabolic syndrome (MetS) in the Peruvian birth cohort and their documented association with MetS components[1]**

| Analyte | Mean | Std. Dev. | Min. | Max. | Available values (%) | Associations with MetS components | Reference |
|---|---|---|---|---|---|---|---|
| **Gastric Inhibitory Polypeptide (GIP)** | 360.5 | 270.0 | 1.8 | 1,365.2 | 213 (98.2) | IR+, BMI+ | (12) |
| **Glucagon-like Peptide 1 (GLP-1) Active/Total** | 3.1 | 10.5 | 0.0 | 129.0 | 30 (13.8) | HDL+, TG-, IR- | (13) |
| **Glucagon** | 35.4 | 27.2 | 2.5 | 207.3 | 90 (41.5) | IR+ | (14) |
| **Interferon gamma (IFN-γ)** | 1.3 | 3.7 | 0.0 | 23.9 | 19 (8.8) | IR+ | (15) |
| **Interleukin 1 beta (IL-1β)** | 0.2 | 0.7 | 0.0 | 4.4 | 9 (4.1) | IR+ | (16) |
| **Interleukin 4 (IL-4)** | 1.1 | 4.5 | 0.0 | 44.0 | 15 (6.9) | IR-, BMI- | (17) |
| **Interleukin 6 (IL-6)** | 5.7 | 34.7 | 0.0 | 428.5 | 45 (20.7) | IR+, HDL- | (18) |
| **Interleukin 10 (IL-10)** | 0.8 | 8.2 | 0.0 | 114.2 | 4 (1.8) | HDL+, TG-, IR- | (19) |
| **Interleukin 17A (IL-17A)** | 0.1 | 0.9 | 0.0 | 8.0 | 3 (1.4) | IR+, BMI+ | (20) |
| **Interleukin 22 (IL-22)** | 1.8 | 9.6 | 0.0 | 76.0 | 8 (3.7) | IR- | (21) |
| **Interleukin 23 (IL-23)** | 1.7 | 15.3 | 0.0 | 187.4 | 2 (0.9) | BMI- | (22) |
| **Interleukin 33 (IL-33)** | 13.5 | 21.8 | 0.0 | 165.1 | 75 (34.6) | IR-, BMI- | (23) |
| **Insulin** | 415.0 | 445.9 | 27.5 | 2,613.2 | 145 (66.8) | IR+, BP+ | (24) |
| **Leptin** | 484.5 | 1,141.1 | 4.7 | 10,303.7 | 135 (62.2) | IR+, BMI+, BP+ | (25) |
| **Monocyte chemoattractant protein-1 (MCP-1)** | 122.8 | 95.4 | 10.9 | 632.8 | 215 (99.1) | HDL-, TG+, IR+, BMI+, BP+ | (26) |
| **Omentin-1** | 1,485,204.9 | 939,705.1 | 129,211.8 | 5,823,200.0 | 217 (100.0) | IR-, BMI-, WC- | (27) |

**Table S1: Summary statistics of candidate outcome biomarkers of metabolic syndrome (MetS) in the Peruvian birth cohort and their documented association with MetS components[1]**

| Analyte | Mean | Std. Dev. | Min. | Max. | Available values (%) | Associations with MetS components | Reference |
|---|---|---|---|---|---|---|---|
| **Pentraxin 3 (PTX3)** | 6,133.0 | 8,789.9 | 396.4 | 51,220.4 | 216 (99.5) | HDL-, TG+, IR+ | (28) |
| **Paraoxonase-1 (PON1)** | 5,168,859.6 | 3,072,601.7 | 9,927.5 | 21,358,020.0 | 210 (96.8) | HDL+, TG-, IR-, BMI- | (29) |
| **Pancreatic polypeptide (PP)** | 338.1 | 274.2 | 11.7 | 1,261.1 | 196 (90.3) | BMI-, IR- | (30) |
| **Peptide tyrosine tyrosine (PYY)** | 109.3 | 83.2 | 10.3 | 561.9 | 20 (9.2) | BMI-, IR- | (31) |
| **Soluble CD40-ligand (sCD40L)** | 259.3 | 462.8 | 0.4 | 3,423.2 | 199 (91.7) | IR+, BMI+, WC+ | (32) |
| **Soluble Leptin Receptor (sOB-R)** | 36,179.3 | 14,389.5 | 7,396.1 | 78,243.8 | 217 (100.0) | HDL+, IR-, WC- | (33) |
| **Tumor Necrosis Factor Alpha (TNF-α)** | 12.9 | 5.9 | 2.8 | 33.6 | 216 (99.5) | IR+, BMI+, WC+, TG+, BP+, HDL- | (34) |
| **Visceral adipose tissue-derived serpin (Vaspin)** | 241.3 | 487.7 | 14.4 | 2,746.9 | 193 (88.9) | IR+, BMI+, TG+ | (35) |

**Table S2: Definitions and distributions of early-life enteric infection and enteropathy markers in the Peruvian birth cohort**

| Variable | Definition | Mean (Std. Dev) |
|---|---|---|
| **Exposures** | | |
| Adiponectin | Within-subject average of plasma concentrations at 7, 15 and 24 months (ug/mL) | 9.3 (2.6) |
| Ferritin (FRTN) | Within-subject average of plasma concentrations at 7, 15 and 24 months (ng/mL) | 32.2 (27.2) |
| Interleukin-8 (IL-8) | Within-subject average of plasma concentrations at 7, 15 and 24 months (pg/mL) | 13.1 (9.0) |
| Proline | Within-subject average of plasma concentrations at 7, 15 and 24 months (μM) | 294.1 (87.9) |
| Serum Amyloid P-Component (SAP) | Within-subject average of plasma concentrations at 7, 15 and 24 months (ug/mL) | 7.3 (2.1) |
| Serotransferrin (Transferrin) | Within-subject average of plasma concentrations at 7 and 15 months (mg/dl) | 312.4 (71.3) |
| Tryptophan | Within-subject average of plasma concentrations at 7, 15 and 24 months (umol/L) | 52.4 (9.6) |
| Alpha-1-Antitrypsin (AAT) | Within-subject average of fecal concentrations, 0-24 months (mg/g) | 0.6 (0.2) |
| Myeloperoxidase (MPO) | Within-subject average of fecal concentrations, 0-24 months (mg/g) | 10,982.4 (3,655.4) |
| Neopterin (NEO) | Within-subject average of fecal concentrations, 0-24 months (nmol/L) | 2,484.7 (835.3) |
| Lactulose % | Within-subject average of percent urinary lactulose recovery at 3, 6, 9, 15 and 24 months | 0.3 (0.2) |
| Mannitol % | Within-subject average of percent urinary mannitol recovery at 3, 6, 9, 15 and 24 months | 2.8 (1.3) |
| Lactulose:mannitol (LM) Z-score | Within-subject average of urinary LM ratio-for-age Z-score at 3, 6, 9 and 15 months | 0.7 (0.4) |
| LAZ-score | Within-subject average of length-for-age Z-score, 0-24 months | -1.6 (0.8) |
| WAZ-score | Within-subject average of weight-for-age Z-score, 0-24 months | -0.5 (0.9) |
| Birth weight | Weight at birth (kg) | 3.1 (0.4) |
| Viral infections | Within-subject total discrete enteric viral infections, 0-24 months | 15.7 (6.2) |
| Bacterial infections | Within-subject total discrete enteric bacterial infections, 0-24 months | 23.1 (7.1) |
| Protozoal infections | Within-subject total discrete enteric protozoal infections, 0-24 months | 8.1 (3.9) |
| Diarrhea episodes | Within-subject total discrete diarrheal episodes, 0-24 months | 10.8 (7.1) |
| **Covariates** | | |
| Sex | Subject's sex | 43.8% male |
| Household income | Average monthly income for subject's household (soles) | 135.3 (52.0) |
| Age at follow-up | Age in months at outcome blood sample collection | 52.6 (13.7) |

Early-life enteric infection and enteropathy markers are associated with changes in adipokine, apolipoprotein and cytokine profiles later in childhood consistent with those of an adverse cardiometabolic disease risk profile in a Peruvian birth cohort.

10. Pang J, Nguyen VT, Rhodes DH, Sullivan ME, Braunschweig C, Fantuzzi G. Relationship of galectin-3 with obesity, IL-6, and CRP in women. J Endocrinol Invest

[Internet]. 2016 Dec 1 [cited 2020 Dec 22];39(12):1435–43. Available from: https://pubmed.ncbi.nlm.nih.gov/27444618/

Early-life enteric infection and enteropathy markers are associated with changes in adipokine, apolipoprotein and cytokine profiles later in childhood consistent with those of an adverse cardiometabolic disease risk profile in a Peruvian birth cohort.

Early-life enteric infection and enteropathy markers are associated with changes in adipokine, apolipoprotein and cytokine profiles later in childhood consistent with those of an adverse cardiometabolic disease risk profile in a Peruvian birth cohort.
